# Prevalence and Genotype Distribution of Human Papilloma Virus (HPV) among Women in Karachi, Pakistan

**DOI:** 10.1101/2025.06.20.25329989

**Authors:** S. Maria Jilani, Angelika Iftner, Hana’a Iqbal, Atia-tul-Wahab, Sharmeen Fayyaz, Thomas Iftner, M. Iqbal Choudhary

**Author notes:** Correspondence: M.I.C; S.F. T.I. These authors contributed equally (S. Maria Jilani, Angelika Iftner). **Author Contributions:** Conceptualization, T.I.; Formal analysis, S.F., and T.I.; Funding acquisition, M.I.C.; Investigation, S. M. J., A.I., and H.I.; Methodology, S. M. J., and A.I.; Project administration, S.F. and T.I.; Resources, M.I.C., A.W., and T.I.; Supervision, M.I.C., S.F., and T.I.; Visualization, S.M.J., and A.I.; Writing— original draft, S.F and TI; Writing—review & editing, S.F., T.I., and M.I.C. All authors have read and agreed to the published version of the manuscript.

## Abstract

We performed a population-based study in densely populated areas of the largest city of Pakistan (Karachi) to determine the overall type- and age-specific prevalence of human papillomavirus (HPV) in the setting of an unscreened and non-vaccinated female population between May 2022 and November 2023. Women (n=3,119) were invited to participate in the study from whom a total of 497 women gave consent and provided cervical samples. HPV positivity was determined and specific HPV genotypes were identified using the INNO LiPa Extra II-line probe assay. Total HPV positivity among all age groups was 16.7%. High risk HPV types (groups 1 and 2) were found in 11.9% whereas low risk types and unclassified types were 4.8% in all samples. Fourteen HR types were detected as single infections (8.4%), highest prevalent types are HPV-31 and HPV-53 (each 1.2%) followed by HPV-68 (1.0%), HPV-16 (0.8%), HPV-33, -39 and -51 (each 0.6%). While HPV-35, HPV-52, HPV-70, HPV-73 and HPV-82 were the least prevalent types. The most prevalent (1.2%) low-risk HPV type detected was HPV-6 of all samples. The highest HPV prevalence (21.3%) was observed in subjects aged 25–34 years (n=220), whereas in the age group above 54 years (n=24) we detected HPV in 12.5% of samples tested. Single HR HPV infections were observed in 8.4%, while multiple HR infections were detected in 4.02% of the 497 women tested. Extrapolating this data to the total female population of Pakistan allows to estimate that about 20 million women are HPV positive. This rough estimation forms a strong basis of an organized cervical cancer screening program using high precision HPV tests for early detection of HPV infections and related diseases including cervical cancer. In addition, this also establish the need of implementation of immunization program with the recently licensed nonavalent human papillomavirus vaccine. This can significantly reduce the future morbidity and mortality from cervical and oral cancer, pre-cancerous lesions and other HPV-related cancers in the female and male population of Pakistan. The current study, therefore, provide a credible basis of further research, and follow-up action on HPV-related disease burden.

## Introduction

Cervical cancer is the 4^th^ most common cancer in women worldwide and the 2^nd^ most common cancer in women of reproductive age. With 662301 new cases and 348874 deaths worldwide in 2022, cervical cancer is a global health challenge (Li, *et al*., 2025; https://gco.iarc.fr/). It is particularly prevalent in developing countries, where about 85% of cervical cancer cases are diagnosed. In many developed countries, the incidence of cervical cancer has declined in recent decades, following the introduction of organized cervical cancer screening programs using cervical cytology (Vaccarella, Franceschi *et al*. 2014). Cervical cytology-based screening programs were launched by developed countries as early as 1970 leading to a reduced incidence of cervical cancer. The incorporation of HPV molecular testing into cervical cancer screening programs further reduced cancers and high-grade cervical intraepithelial neoplasia (Corrêa, *et al*., 2022; Liu *et al*., 2024; Ronco, Dillner *et al*. 2014). This approach has been adopted by only a few developing countries, because of a lack of organized screening the incidence of cervical cancer remains high in developing countries (Bruni *et al*., 2022; Cohen *et al*., 2019).

Persistent infection with HR-HPV types has been established as a necessary causal step in the development of cervical cancer. Approximately 30 HPV types infect the genital mucosa, particularly in the anogenital tract of women and men (Brautigam, *et al*., 2022). According to their carcinogenic potential, HPVs are divided into high and low risk types. low-risk (LR) types mainly cause warts in the external genital tract (e.g. condylomata acuminata) and low-grade cervical intraepithelial neoplasia (CIN1) at the cervix. Whereas, high-risk (HR) HPVs are associated with the development of cervical, vulvar and anal leisons, and cancers. HPV types 16, 18, 31, 33, 35, 39, 45, 51, 52, 56, 59 and 68 are classified as group I/IIA high risk (HR), and types 26, 53, 66, 67, 69, 70, 73, 82, 85, 34, and 30 are group IIB carcinogens or potential high risk. While HPV types 6, 11, 40, 44, 62, 81, and 54 are classified as low risk (LR) with low evidence of carcinogenicity. All other types affecting the anogenital tract have not yet been classified (Wei *et al*., 2024.)

Pakistan is the 5^th^ most populous country in the world, where incidence of cancer is higher than its neighboring countries. Cervical cancer accounts for about 2.3% of all cancer-related deaths among women in the country. According to the GLOBOCAN 2024 report, there were approximately 4,762 new cervical cancer cases and 3,069 related deaths in Pakistan in 2022 (GLOBOCAN, 2024). Primary cause of cervical cancer is persistent HPV infection.

In Pakistan, the high prevalence of cervical cancer is largely due to late diagnosis and lack of vaccination programes. Women are unaware of the risk factors of cervical cancer, screening possibilities, and HPV testing. In addition, sexual practices in Pakistan differ from those in Western countries, and any discussion of sexual behavior, including sexually transmitted infections, is a social taboo. These are the major challenges in raising awareness about HPV infections and related cancers. Therefore, it is crucial to conduct epidemiological studies to assess the prevalence of HPV in Pakistani women, and to lay the foundation for future cervical cancer screening programs.

In current study we used the sensitive INNO Lipa Extra II line probe assay, based on the principle of reverse hybridization to estimate the prevalence of HPV infections in women from Karachi, Pakistan. The highly sensitive INNO Lipa test identify the full spectrum of single and mixed genital HPV infections (32 genotypes) was deployed to obtain the full picture of HPV infection. A questionnaire to collect demographic and behavioral data on potential risk factors for HPV positivity was also used. A rather high prevalence of HPV (16.7%) in women aged between 25-60 years was observed. Volunteers were recruited among women seeking medical care in tertiary hospitals for various reasons.

## Material and methods

### Study population and recruitment

This study was carried out in collaboration with the Gynecology departments of different hospitals. Recruitment was carried out from May 2022 to November 2023. Hospitals of Central, South, and West districts of Karachi, and geographical clusters including Malir, Landhi, DHA, and HUB (at community level/municipalities), were contacted for recruitment of participants from each of these clusters after taking consent.

### Ethical Approval

The independent ethics committee of International Center for Chemical and Biological Sciences (ICCBS) approved the study protocol (Study# 076-PS-2022) as ICCBS/IEC-076-PS-2022/Protocol/1.0.

### Sample collection

Pap smears samples were collected from women who visited an outpatient gynecology department. In total 9 hospitals were invited to participate in the study. Of these, 4 gynecologists from 5 hospitals agreed to participate. The participants were briefed about the aims and objectives of the research, and informed consent froms were obtained before sample collection. Participants or concerned doctors were asked to fill a questionnaire. The questionnaire was divided into four sections: (1) general demographic information and (2) information regarding cancer screening and history (3) information on sexual behavior and smoking habits, and (4) information on previous knowledge or awareness about HPV screening.

### HPV Testing

Pap smears were collected and transported to the National Institute of Virology (PCMD) at the ICCBS, University of Karachi (NIV) at ambient temperature in viral transfer medium (Sansure Biotech, China), and processed for DNA extraction either on the same day or stored at 4°C until processed.

### DNA Extraction

DNA was extracted using QIA amp DNA Mini Kit as per manufacturer instructions (Qiagen QIA amp kit, Germany), and stored at −20°C until use. Samples were shortly spun before extracting DNA to dislodge the cells from cytobrush. A total volume of 500 μL of the sample was lysed with 180 μL of lysis buffer in the presence of proteinase K. The resultant mixture was vortexed for 15 seconds and incubated at 56°C for 35 minutes, and resuspended after addition of buffer 2 at 70°C for 10 minutes. Tubes were then centrifuged to pellet down the extracted DNA. Supernatant was discarded, and DNA pellet was washed. Purified DNA was stored at -20°C until further use.

### HPV Testing and genotyping

INNO-LiPA HPV Genotyping Extra II (Fujirebio) was used to test the presence of HPV DNA. Testing was carried out according to the manufacturer’s protocol. INNO-LiPA identifies 32 HPV genotypes of which 13 are defined as high-risk (HR) HPV types (HPV16/18/31/33/35/39/45/51/52/56/58/59/68), and the rest (6/11/26/40/42/43/44/ 53/54/61/62/67/66/70/73/81/82/83/89) were classified as low-risk, potentially high risk or unclassified types. Samples testing positive for HPV but without identifying a specific genotype were classified as HPV X. Results were communicated to concerned gynecologist who were responsible for forwarding results to the participants.

## Results

A total of n=3119 women were invited to participate in the study (Fig. 1A), among them 497 women consented and allowed to collect samples. Characteristics of the cohort are shown in Table 1.

**Figure 1:**
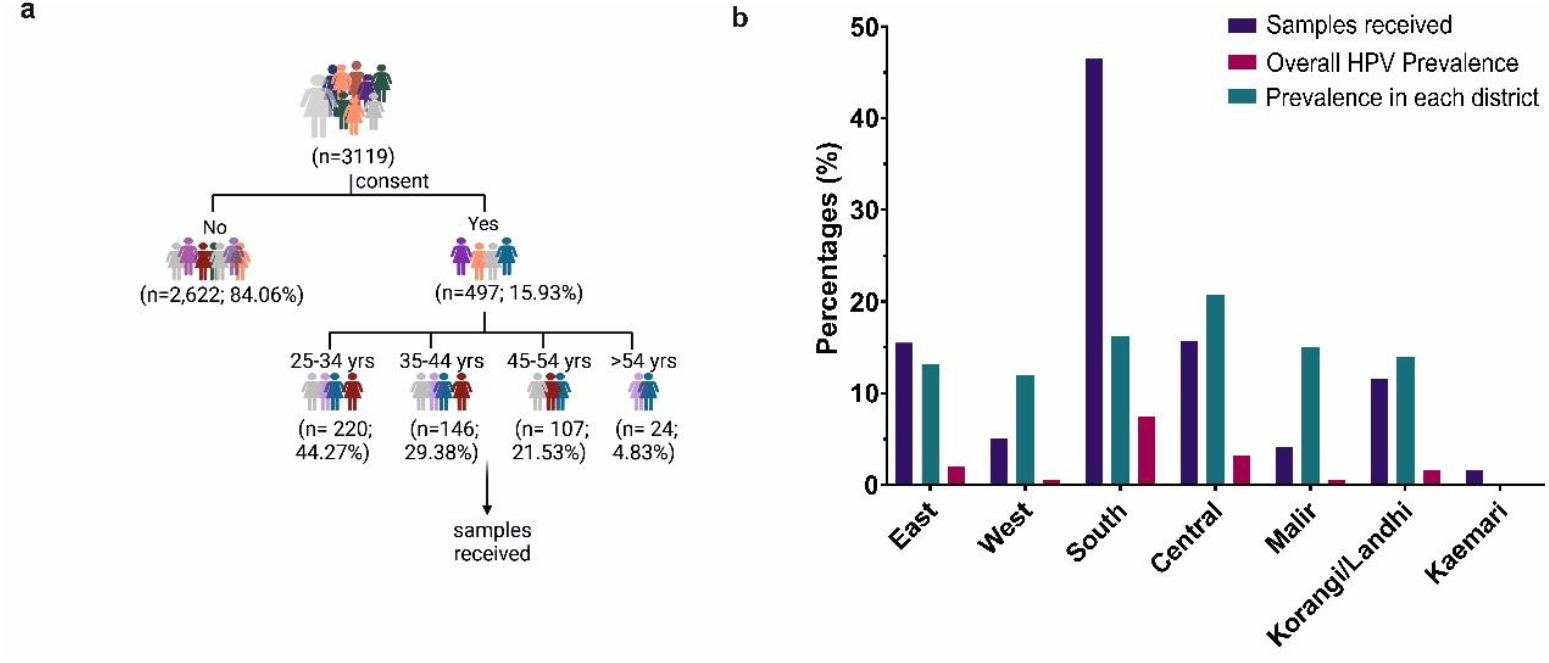
Inclusion and distribution of study participants from different areas of Karachi. (a) Scheme of participant recruitment. After receiving a consent for inclusion in study each participant was further classified in age-specific group. (b) Comparison of overall HPV prevalence in different areas of Karachi as a ratio of the number of samples received from each area.

**Table 1.** Characteristics of the study participants.

|  | Total<br>n = 497<br>n (%) | HPV-negative<br>n= 414<br>n (%) | HPV-positive<br>n = 83<br>n (%) |
| --- | --- | --- | --- |
| **Region/District of Karachi** |  |  |  |
| East | 78 (15.7) | 66 (15.9) | 12 (14.4) |
| West (Orangi) | 25 (5.03) | 22 (5.31) | 3 (3.6) |
| South | 232 (46.7) | 191 (46.13) | 41 (49.4) |
| Central | 78 (15.7) | 61 (14.7) | 17 (20.5) |
| Malir | 20 (4.02) | 17 (4.10) | 3 (3.6) |
| Korangi/landhi | 57 (11.47) | 49 (11.8) | 8 (9.6) |
| Kaemari | 08 (1.61) | 08 (1.93) | 0 |
| **Annual Income** |  |  |  |
| <2,000 USD | 147 (29.57) | 125 (30.2) | 22 (26.5) |
| 2,000–6,000 USD | 157 (31.6) | 135 (32.6) | 22 (26.5) |
| >24,000 USD | 186 (37.4) | 154 (37.2) | 32 (38.5) |
| Missing | 07 (1.41) | 00 | 07 (8.4) |
| **Smoking** |  |  |  |
| No | 398 (80.1) | 334 (80.67) | 64 (77.1) |
| Yes | 99 (19.9) | 80 (19.3) | 19 (22.9) |
| **PAP test awareness** |  |  |  |
| Yes | 54 (10.8) | 46 (11.11) | 08 (9.6) |
| No | 443 (89.1) | 368 (88.9) | 75 (90.3) |
| **Contraceptive Practice** |  |  |  |
| No | 338 (68.0) | 284 (68.6) | 54 (65.0) |
| Intrauterine devices (IUD) | 90 (18.0) | 76 (18.3) | 14 (16.8) |
| Oral | 47 (9.4) | 37 (8.9) | 10 (12.0) |
| Condoms | 21 (4.2) | 17 (4.1) | 04 (4.8) |
| Missing | 01 (0.2) | -- | 01 (1.2) |
| **Clinical symptoms\*** |  |  |  |
| Yes | 352 (70.8) | 291 (70.3) | 61 (73.5) |
| No | 145 (29.17) | 123 (29.7) | 22 (26.5) |

\*Clinical symptoms include unusual vaginal discharge, postmenopausal or post coital bleeding, and irritation or discomfort.

### Prevalence of HPV infections

HPV positivity and specific HPV genotypes were identified using the INNO LiPa Extra II-line probe assay. Total HPV positivity among all age groups was 16.7%. High risk HPV types (groups 1 and 2) were found in 11.9% whereas low risk and unclassified types were in 4.8% of all women (table-2; Fig 2a). Fourteen HR types were detected as single infections (8.4%) with HPV-31 and -53 (each 1.2%) followed by HPV-68 (1.0%), HPV-16 (0.8%), HPV-33, -39, and -51 (each 0.6%) while HPV-35, HPV-52, HPV-70, HPV-73 and HPV-82 were the least prevalent types (table-3). The most prevalent low-risk HPV identified was HPV-6 in 1.2% of all samples. The highest HPV prevalence of 21.3% was observed in subjects aged 25–34 years, while in the age group above 54 years, HPV prevalence was 12.5% (Fig. 2b).

**Table 2.**
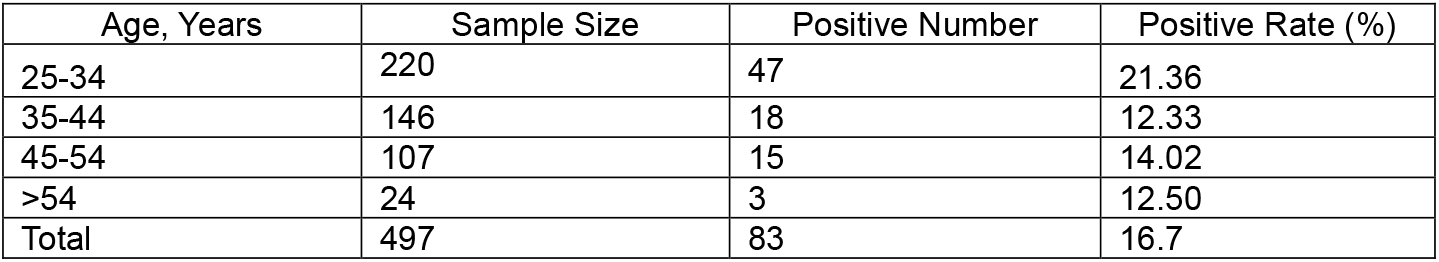
HPV Prevalence of the Study Participants.

| Age, Years | Sample Size | Positive Number | Positive Rate (%) |
| --- | --- | --- | --- |
| 25-34 | 220 | 47 | 21.36 |
| 35-44 | 146 | 18 | 12.33 |
| 45-54 | 107 | 15 | 14.02 |
| >54 | 24 | 3 | 12.50 |
| Total | 497 | 83 | 16.7 |

**Table 3.**
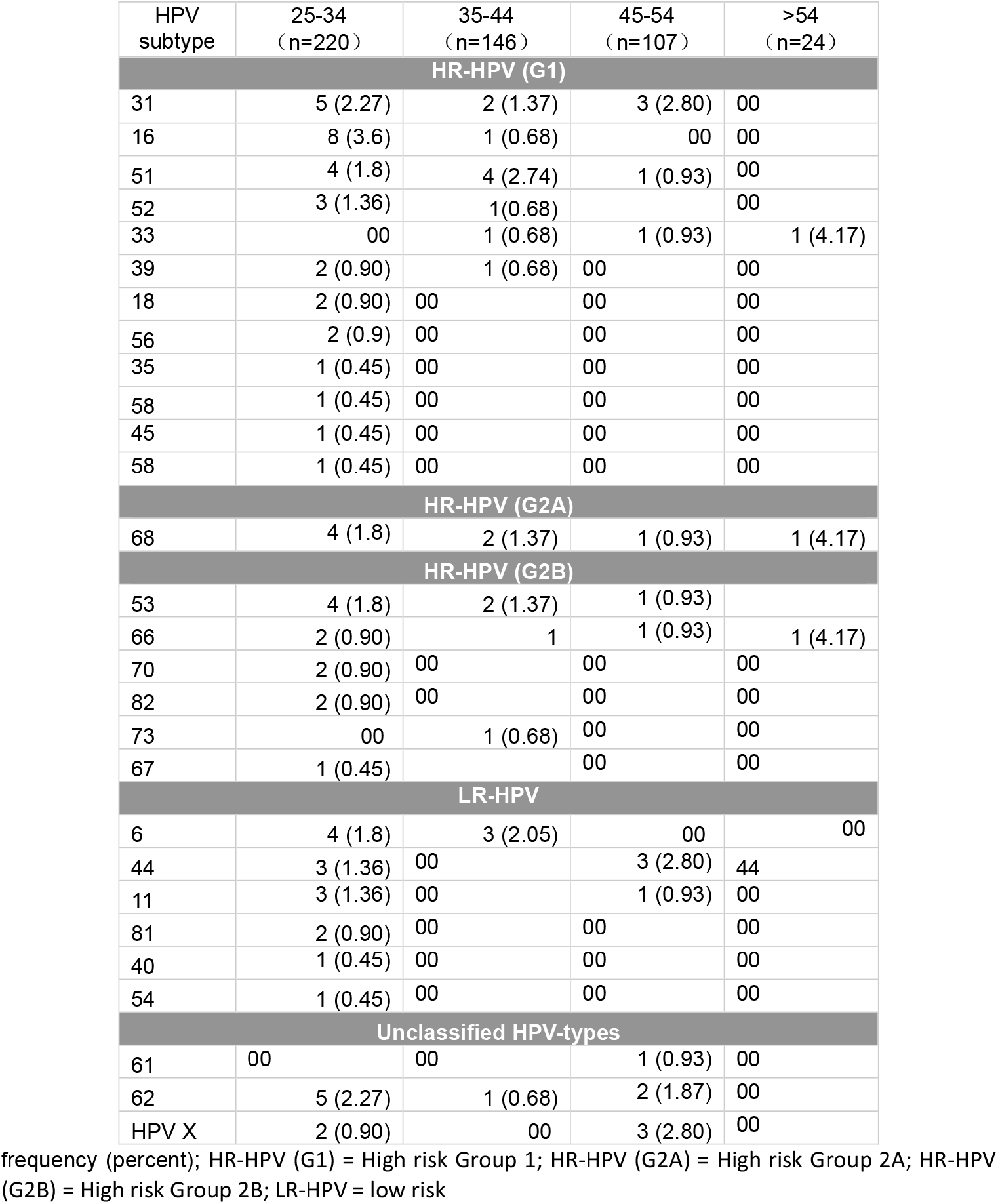
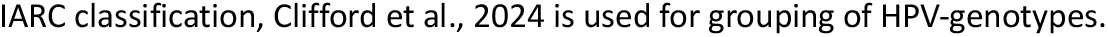
Infection with high and low risk, and unidentified-HPV subtypes in different age groups.

| HPV subtype | 25-34<br>(n=220) | 35-44<br>(n=146) | 45-54<br>(n=107) | >54<br>(n=24) |
| --- | --- | --- | --- | --- |
| **HR-HPV (G1)** |  |  |  |  |
| 31 | 5 (2.27) | 2 (1.37) | 3 (2.80) | 00 |
| 16 | 8 (3.6) | 1 (0.68) | 00 | 00 |
| 51 | 4 (1.8) | 4 (2.74) | 1 (0.93) | 00 |
| 52 | 3 (1.36) | 1(0.68) |  | 00 |
| 33 | 00 | 1 (0.68) | 1 (0.93) | 1 (4.17) |
| 39 | 2 (0.90) | 1 (0.68) | 00 | 00 |
| 18 | 2 (0.90) | 00 | 00 | 00 |
| 56 | 2 (0.9) | 00 | 00 | 00 |
| 35 | 1 (0.45) | 00 | 00 | 00 |
| 58 | 1 (0.45) | 00 | 00 | 00 |
| 45 | 1 (0.45) | 00 | 00 | 00 |
| 58 | 1 (0.45) | 00 | 00 | 00 |
| **HR-HPV (G2A)** |  |  |  |  |
| 68 | 4 (1.8) | 2 (1.37) | 1 (0.93) | 1 (4.17) |
| **HR-HPV (G2B)** |  |  |  |  |
| 53 | 4 (1.8) | 2 (1.37) | 1 (0.93) |  |
| 66 | 2 (0.90) | 1 | 1 (0.93) | 1 (4.17) |
| 70 | 2 (0.90) | 00 | 00 | 00 |
| 82 | 2 (0.90) | 00 | 00 | 00 |
| 73 | 00 | 1 (0.68) | 00 | 00 |
| 67 | 1 (0.45) |  | 00 | 00 |
| **LR-HPV** |  |  |  |  |
| 6 | 4 (1.8) | 3 (2.05) | 00 | 00 |
| 44 | 3 (1.36) | 00 | 3 (2.80) | 44 |
| 11 | 3 (1.36) | 00 | 1 (0.93) | 00 |
| 81 | 2 (0.90) | 00 | 00 | 00 |
| 40 | 1 (0.45) | 00 | 00 | 00 |
| 54 | 1 (0.45) | 00 | 00 | 00 |
| **Unclassified HPV-types** |  |  |  |  |
| 61 | 00 | 00 | 1 (0.93) | 00 |
| 62 | 5 (2.27) | 1 (0.68) | 2 (1.87) | 00 |
| HPV X | 2 (0.90) | 00 | 3 (2.80) | 00 |

frequency (percent); HR-HPV (G1) = High risk Group 1; HR-HPV (G2A) = High risk Group 2A; HR-HPV (G2B) = High risk Group 2B; LR-HPV = low risk
IARC classification, Clifford et al., 2024 is used for grouping of HPV-genotypes.

Single HR-HPV infections were observed in 8.4%, while multiple HR infections were present in 6.84% of the 497 women tested (table-4). The HR-HPV genotypes, HPV31 (2.04%, n=10), HPV 16 and HPV 51 (1.8%, n=9) were most common. It is worth noting that HPV-45, -52, -54, -58, -59, and -67 were found only in multiple infections with other HPV types. No cases of multiple infections were found in women aged above 54 years, as compared to the age group of 25-34 where multiple infections found in 15 samples (6.82%) (Tables S1-S2).

**Figure 2.**
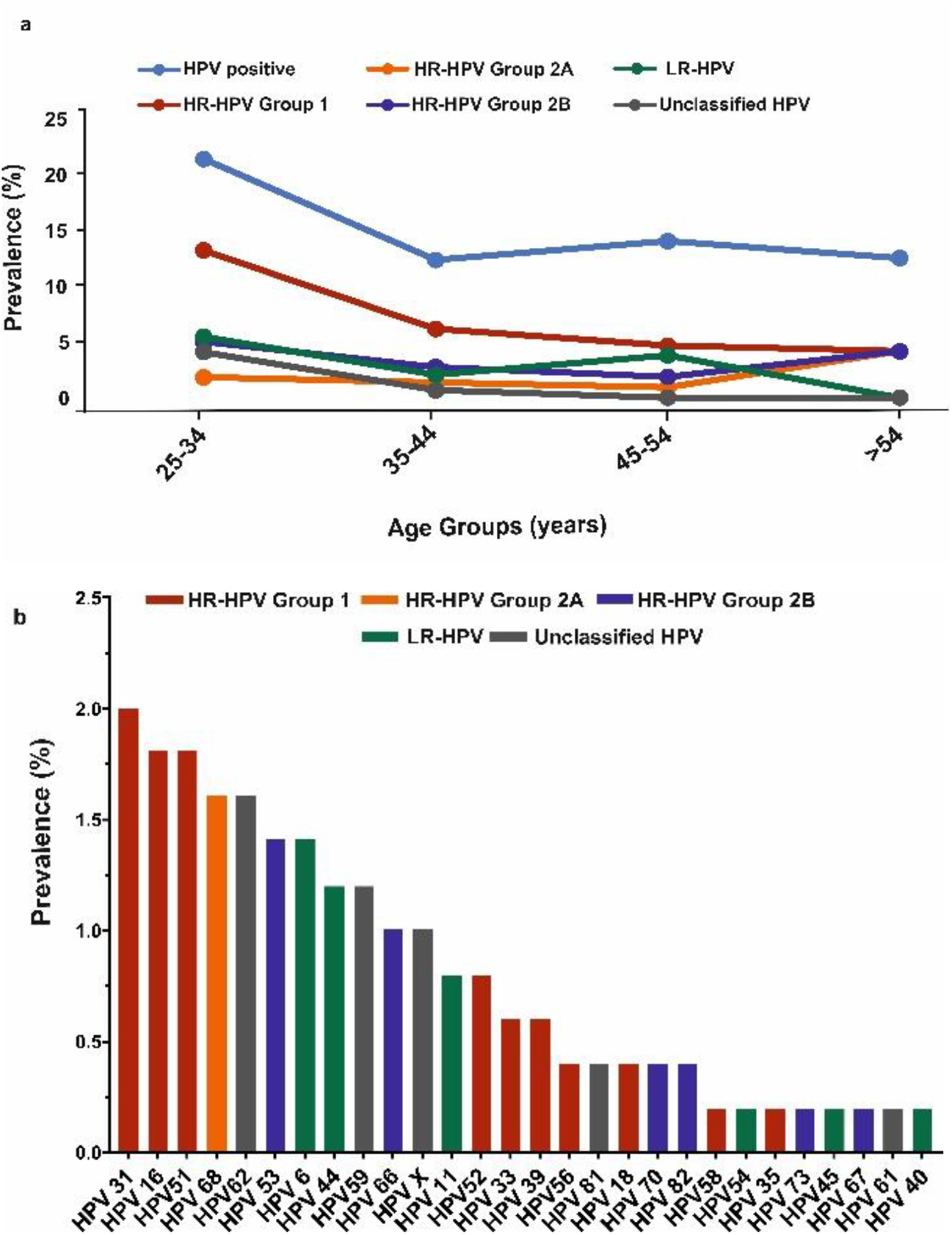
Prevalence of HPV infections. (a) Percent prevalence of HPV infections with low-and high-risk HPV types in different age groups (b) Type-specific HPV prevalence for participants recruited in the study. High risk-HPV (HR-HPV) groups 1 (carcinogenic), 2A (probably carcinogenic), and 2B (potential carcinogenic) are designated according to IARC classification.

**Table 4.**
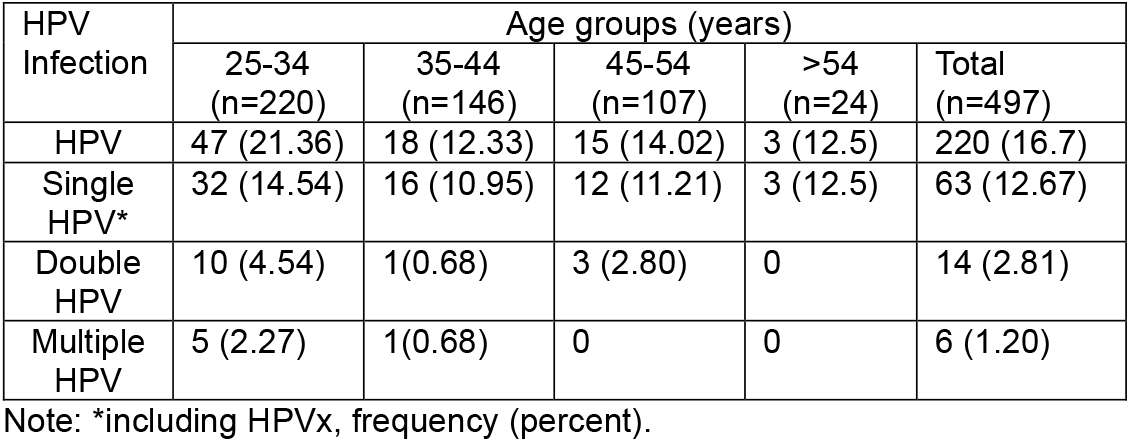
Distribution of single, double, and multiple HPV infections in study groups.

| HPV Infection | Age groups (years) |  |  |  |  |
| --- | --- | --- | --- | --- | --- |
|  | 25-34<br>(n=220) | 35-44<br>(n=146) | 45-54<br>(n=107) | >54<br>(n=24) | Total<br>(n=497) |
| HPV | 47 (21.36) | 18 (12.33) | 15 (14.02) | 3 (12.5) | 220 (16.7) |
| Single HPV* | 32 (14.54) | 16 (10.95) | 12 (11.21) | 3 (12.5) | 63 (12.67) |
| Double HPV | 10 (4.54) | 1(0.68) | 3 (2.80) | 0 | 14 (2.81) |
| Multiple HPV | 5 (2.27) | 1(0.68) | 0 | 0 | 6 (1.20) |

Note: \*including HPVx, frequency (percent).

### HPV Genotype distribution in women across different age groups

Prevalence of HPV infections in different age groups is presented in table-3. Women were categorized into four age groups, 25–34 years, 35-44 years, 45–55 years, and above 54 years.

It was interesting to note that infections with LR-HPV also decreased with increasing age, no infection with LR-HPV types were found in women aged above 54 years. whereas the prevalence of LR-HPV was 3.2% (n=7) in the age group of 25-34 years. High-risk HPV-16 (n=8; 3.6%), and HPV-31 (n=5; 2.3%), were the most prevalent in the women aged 25-34 years, followed by HPV-51, -68, -53 (N=4; 1.8%, each) in age groups of 25-34 years in single and multiple infections grouped together.

### 3.1 Demographic characteristics of participants

Persistent or acute HPV infection depend on certain social and cultural practices. It is, therefore, important to consider demographic factors in the epidemiology of HPV infections. Known risk factors include age, early sexual debut, number of sex partners, smoking, etc. were evaluated with the help of a questionnaire. Age of the participants ranged from 25 to 60 years, with a mean age of 37 years. The participants were then sorted into four age groups, 25 to 34 years (n =220, 44.3%), 35 to 44 years (n= 146, 29.4%), 45 to 54 years (n=107, 21.5%), and >54 years (n= 24, 4.8%). The participants registered were residents of Karachi with an exception of 10 participants (2.04%) that were from Quetta (Baluchistan) and differed in region of residence, educational status, and socioeconomic background. Karachi is the 12^th^ largest cities of the world and the largest in Pakistan with over 20 million population. Karachi’s demographics is characterized by immigration from all regions of Pakistan. Study participants were recruited from all districts of Karachi. Largest number of participants were recruited from district South (n= 228, 46.5%), followed by East and Central (n=76, 15.5%), and Korangi/Landhi districts (11.6%). While 2-5% of women were from Malir (n=20, 4.08%), Kaemari (n=08, 1.6%), and Karachi West (n=25, 5.1%). A prevalence of 12.0-20% was found in different districts, highest in district Central (Figure-1b).

Socioeconomic background is an established factor affecting the HPV prevalence as it can define access to resources, awareness, and hygienic practices (Avni-Singer *et al*., 2020; Harling, *et al*., 2013). Interestingly, we did not observe any effect of annual income on the HPV prevalence (table-1). One of the main reasons could be that 46.5% of women were recruited from district South, having similar financial, educational and social status. A recent study conducted in Brazil to find the impact of socioeconomic status on HPV infection among young women, demonstrated that the prevalence of overall HPV was similar among participants from different the social classes (Kops, Horvath *et al*. 2021). This highlights the importance of screening and provision of vaccines to the entire population.

### Risk factors for HPV infection

Major risk factors which are associated with cervical HPV infections include marital status, smoking habits, and multiple sexual partners. Notably, only 2 women had multiple partners and both were found positive for high-risk HPV types.

HPV infections peaked in young women around initial years of sexual debut and declined in the higher age groups. This is in line with the other studies showing highest HPV prevalence among younger women for all regions of the world, reflecting a higher probability of acquiring new infections at the young age.

No differences with regard to HPV prevalence were observed between women using oral contraceptives, intrauterine device (IUD) and condoms. However, in our study groups only a minor fraction of women (n=57, 11.5%) did use any form contraceptives, which is in contrast to studies from the western world. Other variables, like professional status and smoking practices were also assessed but no significant associations with HPV prevalence were observed (table-1).

## Discussion

Using an age-stratified sampling strategy, we have found an overall prevalence of 16.7% in women from different regions of Karachi. It is one of the first study testing HPV infections in general women population. The oncogenic HPV types 31, 16, and 51 were the most prevalent, and HPV-6 was by far the commonest LR type. The highest prevalence of the HR-HPV types was seen among the women in aged 25 to 34 years. Similarly, prevalence of both single and multiple HPV infection peaked among young women.

In a total of three reports present up till now for HPV prevalence from Karachi there is a continuous increase from 2.8% to 15.4% from 2004-2016 (Raza, Franceschi *et al*. 2010, Shahid, Kazmi *et al*. 2015, Khan, Noonari *et al*. 2016). Other studies conducted in Islamabad, Rawalpindi, and Lahore suggested a lower HPV prevalence of 2-5% (Aziz, *et al*., 2018; Minhas, Kashif, *et al*., 2021). This relatively lower rate of infection could be attributed to the differences in detection techniques used and sample size. Studies of HPV prevalence previously showed that differences in sampling sites and HPV DNA testing methods resulted in different estimates (Hebnes *et al*., 2015; Munk *et al*., 2024). Heterogeneity in HPV prevalence has been observed even in comparable study populations. HPV testing for identification of women at higher risk for the development of cervical cancer significantly differs from molecular testing for other medically relevant viruses. Ultimate sensitivity is required for the detection of HPV types that are common in women with latent infection e.g., HPV53 and HPV66, and responsible for low grade precancerous cervical lesions (Poljak *et al*., 2012). We used INNO-LIPA technique, which is highly sensitive for type-specific monitoring of genotypes present in the tested samples. Moreover, Lipa can detect infections with a very low viral load that would not be picked up by a clinically specific HPV test. This is in-line with a study published about Danish man which tested all the samples side-by-side with hybrid capture (HC2) and Lipa and found twice as high prevalence with the Lipa (Hebnes *et al*., 2015).

In most of the developed countries with immunization programs, infections with HR-HPV decreased significantly after introduction of school-based vaccination programs (Steben *et al*., 2018). Whereas, most of the developing countries, China, Korea, and Gulf Cooperation Council (GCC) countries lacking vaccination and cervical cancer screening programs has 15-20% HPV prevalence (Mohammed *et al*., 2019; Ouh *et al*., 2016; Huang *et al*., 2022; Yang *et al*., 2021). As prevalent HPV genotypes may vary significantly in different regions, therefore, specific HPV prevalence data is critically important for screening programs, and vaccine selection. Unfortunately, none of the previous studies present until now give significant information about HPV genotype distribution in Pakistan. We found a high prevalence of HPV 16 and 31, followed by high-risk HPV types 51, and 59 whereas HPV 18 was found only in 2 women (Fig 2b). This finding came as a surprise as HPV genotypes 16, and 18 have been reported to contribute most in the development of cervical cancer (Wang *et al*., 2018; Li *et al*., 2019). Notably, a recent study published for prevalence and distribution of HPV infections in Xiamen, China ranked HPV 18 seventh in HR-HPV types detected (Shen, Huang *et al*., 2023; Huang *et al*., 2022). Thus, HPV genotype distribution varies significantly in different geographical locations. These findings further contribute in the appropriate selection and use of HPV vaccines in targeted high-risk population for the prevention of cervical cancer.

Cervical coinfection with more than one HPV genotype is common. Some studies demonstrated that multiple infections persist for longer duration as compared to a single infection (Rideg, Dergez *et al*. 2022). While some other reports suggest that multiple HPV infections are associated with greater risk of development, and progression of lesions (Trottier, Mahmud *et al*. 2006). To date, there is no consensus whether multiple HPV types occur randomly or through a competitive or cooperative relationships. In our study, the prevalence of single HPV infections was higher than that of double and multiple HPV infections, yet high risk types HPV52, 58, and 59 were detected only in multiple infections. Whether single or multiple infections increase the risk of cervical cancer is yet to be determined and warrants further investigation.

Sexual practices in Pakistan are different from European/Western countries, 99.6% of recruited women were monogamous, and only two women reported to have multiple partners. Although, both of these women were tested positive for HR-HPV we cannot establish the correlation. This is in agreement with studies conducted in Malaysia, Kingdom of Saudi Arabia, and Iran having similar customs, where HPV prevalence is relatively high even among monogamous women and number of sexual partners was not found to be the key factor affecting HPV prevalence (Alsbeih, Al-Harbi *et al*. 2013, Mousa, Al-Amri *et al*. 2019, Sabet, Mosavat *et al*. 2021, Bakhshani, Ganjali *et al*. 2023).

Although this study presents the detailed investigation of HPV infections in general women population of Pakistan, these findings were not combined with cytology or histological results. Moreover, most of the women (46.5%, n=228) who gave consent were from South district, which were relatively more educated. This potential over representation of health-conscious women may mask the actual association between certain risk factors and HPV infections. In this context, it is even more important to disseminate the awareness about HPV testing and cervical cancer screening even among women from marginalized socioeconomic groups.

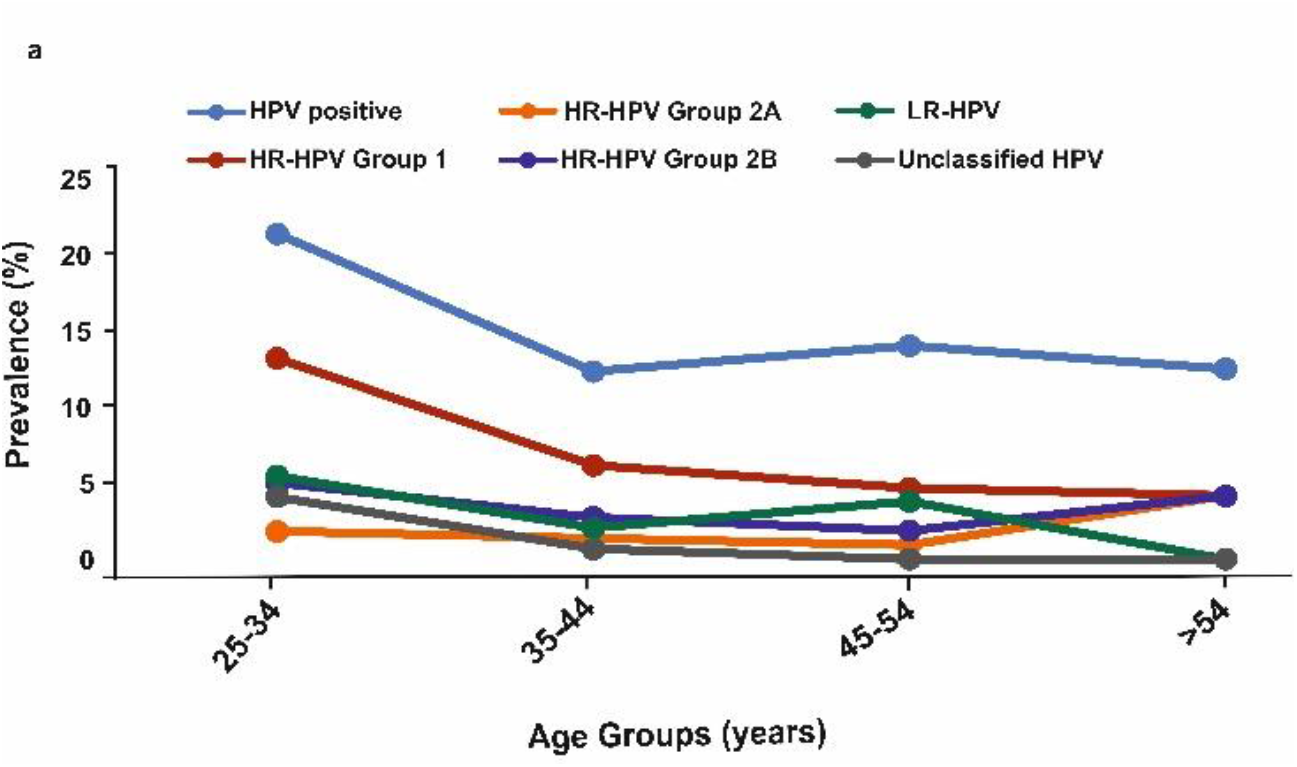

## Conclusion

The population composition of Karachi is progressively complex. Thus, it is important to understand the overall HPV prevalence, and genotype distribution. Among the participants in this study, the overall HPV positive rate, including HR-HPV and LR-HPV was 16.7%. since we used INNOLipa for detection of different HPV types, increased prevalence as compared to previous studies was expected. Most common types identified were HPV 16, 31, and 51 while HPV 6 followed by HPV44, and HPV 11 were the most prominent among LR-HPV types.

Our results emphasize the importance of use of validated HPV tests for HPV vaccine development, and implementation and monitoring of vaccination programs. These findings provide fundamental information for cervical cancer screening, for health ministries to invest in the development of next-generation HPV-targeted vaccination. Nationwide surveys of cervical HPV prevalence and its related risk factors will establish the basis for future assessments of the impact of HPV immunization programs and the dynamics of risk factors for HPV infection in the population of Pakistan.

## Data Availability

All data produced in the present work are contained in the manuscript

