## supplementary material for "Prevalence and Genotype Distribution of Human Papilloma Virus (HPV) among Women in Karachi, Pakistan"

| HPV Genotype | *Single HPV Infection*  n (%) | *Double HPV Infection* n (%) | *Multiple HPV Infection*  n (%) |
| --- | --- | --- | --- |
| HPV 31 | 6 (1.2) | 3 (0.6) | 1 (0.2) |
| HPV 16 | 4 (0.8) | 1 (0.2) | 4 (0.8) |
| HPV51 | 3 (0.6) | 4 (0.8) | 2 (0.4) |
| HPV62 | 5 (1.0) | 1 (0.2) | 1 (0.2) |
| HPV 68 | 5 (1.0) | -- | 2 (0.4) |
| HPV 6 | 6 (1.2) | -- | 1 (0.2) |
| HPV 53 | 6 (1.2) | -- | 1 (0.2) |
| HPV59 | -- | 1 (0.2) | 5 (1.0) |
| HPV 44 | 1 (0.2) | 1 (0.2) | 3 (0.6) |
| HPV 66 | 4 (0.8) | -- | 1 (0.2) |
| HPV52 | -- | 4 (0.8) | - |
| HPV 11 | 1 (0.2) | 1 (0.2) | 1 (0.2) |
| HPV 39 | 3 (0.6) | -- | -- |
| HPV 33 | 3 (0.6) | -- | -- |
| HPV 82 | 2 (0.4) | -- | -- |
| HPV 70 | 1 (0.2) | -- | 1 (0.2) |
| HPV 18 | 2 (0.4) | -- | -- |
| HPV56 | 1 (0.2) | -- | 1 (0.2) |
| HPV 81 | 1 (0.2) | -- | 1 (0.2) |
| HPV 40 | 1 (0.2) | -- | -- |
| HPV54 | -- | -- | 1 (0.2) |
| HPV 61 | 1 (0.2) | -- | -- |
| HPV 67 | -- | -- | 1 (0.2) |
| HPV 73 | 1 (0.2) | -- | -- |
| HPV45 | -- | -- | 1 (0.2) |
| HPV58 | -- | -- | 1 (0.2) |
| HPV 35 | 1 (0.2) | -- | -- |

Table-S1: Overall prevalence of different HPV genotypes in study participants.

Table-S2: Age specific distribution of HPV genotypes as single or multiple infections.

| **HPV Genotype** | ***Single HPV Infection***  ***n (%)*** | ***Double HPV Infection***  ***n (%)*** | ***Multiple HPV Infection n (%)*** |
| --- | --- | --- | --- |
| **25-34 years** | | | |
| HPV 16 | 3 (1.4) | 4 (1.8) | 1 (0.45) |
| HPV 31 | 2 (0.9) | -- | 3 (1.4) |
| HPV59 | -- | 1 (0.2) | 4 (1.8) |
| HPV51 | 1 (0.45) | -- | 3 (1.4) |
| HPV 68 | 1 (0.45) | 3 (0.6) | -- |
| HPV 53 | 3 (1.4) | 1 (0.45) | -- |
| HPV62 | 3 (1.4) | -- | 1 (0.45) |
| HPV 6 | 3 (1.4) | 1 (0.45) | -- |
| HPV52 | -- | -- | 3 (1.4) |
| HPV 44 | -- | 2 (0.9) | 1 (0.45) |
| HPV 11 | 1 (0.45) | 1 (0.45) | 1 (0.45) |
| HPV 70 | 1 (0.45) | 1 (0.45) | -- |
| HPV 82 | 2 (0.9) | -- | -- |
| HPV56 | 1 (0.45) | 1 (0.2) | -- |
| HPV 81 | 1 (0.45) | 1 (0.45) | -- |
| HPV 39 | 2 (0.9) | -- | -- |
| HPV 18 | 2 (0.9) | -- | -- |
| HPV 66 | 2 (0.9) | -- | -- |
| HPV 35 | 1 (0.45) | -- |  |
| HPV 40 | 1 (0.45) | -- | -- |
| HPV54 | -- | 1 (0.45) | -- |
| HPV 67 | -- | 1 (0.45) | -- |
| HPV45 |  | 1 (0.45) | -- |
| HPV58 | -- | -- | 1 (0.45) |
| **35-44 years** | | | |
| HPV51 | 2 (1.4) | 1 (0.7) | 1 (0.7) |
| HPV 6 | 3 (2.0) | -- | -- |
| HPV 31 | 2 (1.4) | -- | -- |
| HPV 68 | 2 (1.4) | -- | -- |
| HPV 53 | 2 (1.4) | -- | -- |
| HPV62 | 1 (0.7) | -- | -- |
| HPV 66 | -- | 1 (0.7) | - |
| HPV 39 | 1 (0.7) | -- | -- |
| HPV52 | -- | -- | 1 (0.7) |
| HPV59 | -- | -- | 1 (0.7) |
| HPV 33 | 1 (0.7) | -- | -- |
| HPV 16 | 1 (0.7) | -- | -- |
| **45-54 years** | | | |
| HPV 44 | 1 (0.93) | 2 (1.9) | -- |
| HPV 31 | 2 (1.9) | 1 (0.93) | -- |
| HPV62 | 1 (0.93) | 1 (0.93) | -- |
| HPV 61 | 1 (0.93) | -- | -- |
| HPV 66 | 1 (0.93) | -- | -- |
| HPV 68 | 1 (0.93) | -- | -- |
| HPV 53 | 1 (0.93) | -- | -- |
| HPV51 | -- | 1 (0.93) | -- |
| HPV 33 | 1 (0.93) | -- | -- |
| **>54 years** | | | |
| HPV 33 | 1 (4.16) | -- | -- |
| HPV 66 | 1 (4.16) | -- | -- |
| HPV 68 | 1 (4.16) | -- | -- |
